# Temporal Cohort Identification for Alzheimer’s Disease with Sequences of Clinical Records

**DOI:** 10.1101/2023.03.03.23286774

**Authors:** Hossein Esitir, Alaleh Azhir, Deborah L Blacker, Christine S Ritchie, Chirag J Patel, Shawn N Murphy

## Abstract

**BACKGROUND:** Alzheimer’s Disease (AD) is a complex clinical phenotype with unprecedented social and economic tolls in an aging global population. Real World Data (RWD) from electronic health records (EHRs) offer opportunities to accelerate precision drug development and scale epidemiological research on AD. A precise characterization of AD cohorts is needed to address the noise abundant in RWD.

**METHODS:** We conducted a retrospective cohort study to develop and test computational models for AD cohort identification using clinical data from 8 Massachusetts healthcare systems. We mined temporal representations from EHR data using a novel transitive sequential pattern mining algorithm (tSPM) to train and validate our models. We then tested our models against a held-out test set from a review of medical records to adjudicate the presence of AD. We trained two classes of models using Gradient Boosting Machine (GBM) to compare the utility of AD diagnosis records versus the tSPM temporal representations (comprising sequences of diagnosis and medication observations) from electronic medical records for characterizing AD cohorts.

**RESULTS:** In a group of 4,985 patients, we identified 219 sequences of medication-diagnosis records for constructing the best classification models. The models with the sequential features improved AD classification by a magnitude of up to 16 percent (over the use of AD diagnosis codes). Six groups of sequences, which we refer to as temporal digital markers, were identified for characterizing the AD cohorts, including sequences that involved (1) a symptom or (2) a risk factor in the past, followed by an AD diagnosis, (3) AD medications, (4) indirect risk factors, symptom management, and potential side effects, (5) comorbidities with possible shared roots or side effects, and (6) plural encounters with of AD diagnosis codes. Discussions of how the identified sequential patterns can be interpreted are provided.

**CONCLUSIONS:** We present sequential patterns of diagnosis and medication codes from electronic medical records, as digital markers of Alzheimer’s Disease. Classification algorithms developed on the sequential patterns can replace standard features from EHRs to enrich phenotype modeling.

## Introduction

Accurate characterization of Alzheimer’s Disease (AD) is complex, often requiring neuro-cognitive, genetic, and imaging markers along with clinical judgment. These markers are seldom routinely collected in clinical care, which limits the scalability and generalizability of these models. Despite the use of detailed biomarkers, prior efforts to create multi-factorial models for AD has often resulted incremental predictive value and external validity, or resulted in inconsistent disease prevalence estimates due to variable case definitions. For example, available models of AD have produced variable and often moderate classification performance, with the area under the receiver operating characteristic (ROC) curves ranging from 0.52 to under 0.86.^1–5^ Further, models developed on neuro-cognitive, genetic, and imaging markers cannot be easily translated to clinical data that are increasingly being used in large epidemiological studies (e.g., insurance claims data) for improving cohort identification.

Suboptimal characterization of AD cohorts can lead to introduction of unnecessary noise, by including a slew of false positive patients to the cohort, and excluding from the cohort those who were falsely identified as negatives. This can, for example, impede the recruitment process for clinical trials aimed at evaluating novel therapies. Further, problematice cohort identification undermines the validity of outcomes research, for instance, that are based on inaccurately designed case and control groups.

Widely available structured clinical data stored in electronic health record (EHR) systems, which include a longitudinal profile of symptoms, diagnoses, medications, and clinical measurements recorded in clinical care, offer possibilities to enrich cohort identification for AD for accurate outcomes research. In addition to the absence of widely accessible detailed specialist evaluation and objective measurements,^6,7^ especially in practices serving underserved areas and populations, AD cases are often poorly documented and misclassified.^8,9^ Providers often assign a non-specific dementia code,^6,7^ and patients sometimes receive conflicting diagnoses.^10^ Further, many cases remain undiagnosed, especially at the early stages of disease presentation due to difficulties in recognizing the signs and symptoms of cognitive impairment during a brief visit.^11^ As a result, the reliability of diagnosis codes for identifying Alzheimer’s Disease patients is suboptimal. The sensitivity and positive predictive value (PPV) of AD diagnosis codes in clinical data range from (sensitivity: 60%-80%) and (PPV: 57%–100%), depending on the clinical data type and diagnosis code.^12,13^

In this study, we adopt a temporal approach for modeling evolving phenotypes with EHR data by Estiri et al.^14–17^ to develop and validate a computational cohort identification algorithm for Alzheimer’s Disease. We demonstrate that classifying true AD cases can be better achieved through computational models that utilize transitive sequences of medications and diagnosis records – rather than AD diagnosis codes – from EHRs. We compare models trained with sequential features of medications and diagnoses to similar algorithms trained using diagnosis codes for AD. Results demonstrate that the models developed with EHR sequences achieve superior classification performances and are consistent with interpretable stories of various sequences of symptoms, recognition, evaluation, and care that often occur. In addition to the hypothesis-generating value of the sequential representations as digital markers of disease, they enable temporal story-telling capabilities and explainable Artificial Intelligence (AI).

## Methodology

We utilized structured medication and diagnosis data for patients who received care at eight healthcare facilities, two tertiary medical centers and four community hospitals, two specialty hospitals, and over 35 primary care centers within the Mass General Brigham (MGB) integrated healthcare system’s footprint in the New England region. Our dataset included patients with samples in the MGB Biobank^18^ until the end of 2020. The use of data for this study was approved by the Mass General Brigham Institutional Review Board (protocol# 2017P000282).

### Study cohort and classification tasks

The study cohort comprised all patients with at least an encounter with an Alzheimer’s Disease or a Dementia diagnosis code. We used diagnosis codes from both the 9th, and 10th revision of International Classification of Diseases (ICD) codes collected under AD and Dementia Phecodes,^19^ which we augmented with historic local codes – the list of ICD codes are available in Table 1S in the Appendix. We identified two classification tasks. First, the goal was to identify patients with a true AD diagnosis, given any diagnosis code (AD, other dementing illness, or nonspecific dementia) – henceforth we call this task and the related study cohort the “Dx AD/Dementia”. Looking at AD diagnosis codes alone, we ran a second classification task on a subset of the first cohort (those with at least 1 specific AD diagnosis code) to identify patients with true AD diagnosis given a specific AD diagnosis code – henceforth, we call this task and the subsequent cohort “Dx AD”.

### Chart reviews for Gold standard labels

To evaluate classification models, we performed chart reviews on 150 patients from our patient pool. In the chart reviews, an expert clinician performed manual review of patients’ charts to adjudicate the presence (or lack thereof) of Alzheimer’s Disease, regardless of the structured diagnosis codes – chart review criteria for adjudicating cases is available in Table 2S in the Appendix.

### Curation of silver-standard labels

To enable semi-supervised learning as described in Estiri *et al*.^20^, we used a Generative Transfer Learning (GTL)^16^ algorithm trained on labeled patient data from other diseases and validated on MGB Biobank data. The GTL algorithms are able to learn transferable healthcare utilization patterns by training on data from other diseases using a small set of disease-agnostic features from EHRs that aim to represent Patients, pRoviders, and their Interactions within the healthcare SysteM (PRISM features) to compute probabilities for a wide range of phenotypes based on their specific recording process. PRISM features and the generative classifiers have exhibited generalizable and transferable distributional characteristics across diseases and patient populations. GTL approach with PRISM classifiers enables the scalable validation of computable phenotypes in EHRs without the need for domain-specific knowledge about specific disease processes.^16^

The model used in this study was a Naive Bayes classifier trained on 101,509 rows of labeled data for 6 PRISM features, including patient gender and numbers of outpatient diagnosis record(s) for the phenotype, unique dates in which the diagnosis codes for the phenotype recorded, unique diagnosis codes for each patient, unique encounters for each patient, and number unique encounters between the first and last phenotype record.

### Feature engineering and modeling

We developed two sets of features. First, we applied the transitive Sequential pattern Mining (tSPM)^14,20^ algorithm to mine sequential representation of medication and diagnosis records from the study cohort’s electronic health records. To mine tSPM representations, the algorithm temporally sorts the medication and diagnosis records from the EHR, selects the earliest record for each clinical observation, and mines all pairs of medication or diagnosis codes that are sequentially related (Figure 1). This results in a large vector of tSPM sequential representation that is then fed into a high-throughput dimensionality reduction algorithm. Second, as the baseline model, we used a list of Alzheimer’s Disease diagnosis ICD-9/10 codes defined in Phecodes. We controlled for age and sex in all models.

**Figure 1.**
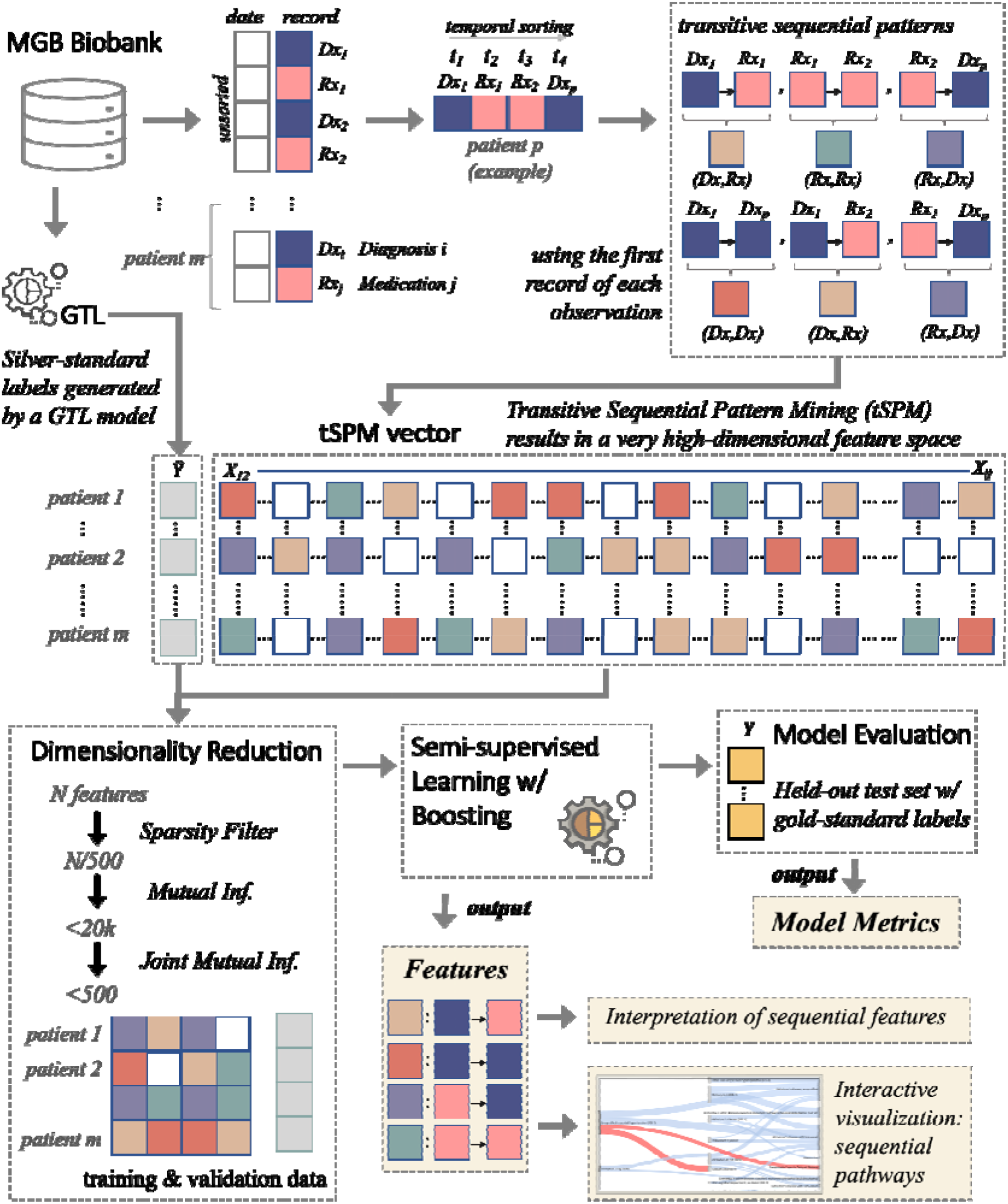
The data pipeline for developing and testing AD classification models through semi-supervised learning with electronic health records data. The pipeline encompasses mining transitive sequential patterns from clinical data, curating silver-standard labels, performing dimensionality reduction, modeling and model evaluations, and interpreting results in an interactive visualization dashboard.

Following Estiri, Vasey, and Murphy (2021), we applied the Minimize Sparsity, Maximize Relevance (MSMR) dimensionality reduction algorithm^14^ to select a small subset of sequential temporal patterns that convey useful information for classification tasks. The resulting sequences were then fed into a Gradient Boosting Machine (GBM),^21^ using the R package gbm.^22^ GBM, through boosting,^23,24^ applies a final step of feature selection to the final modeling features. This model also provides a relative tree-based feature importance metric, determined based on the cumulative use of a given sequential feature in each decision tree step across all trees used in the final boosted model. We used the feature importance score in each classification task to rank the final sequential features.

### Understanding the sequential features

To understand the meaning and functions of the obtained sequences of EHR observations, we used visualization techniques along with clinical expertise. We leveraged the final set of tSPM representations to develop a dashboard that temporally connects the sequences for creating pathway visualization. We also created donut charts to understand the marginal benefit of the sequential approach by comparing the positive predictive values of each elements of a sequence and the full sequence. The positive predictive values are computed based on the true positive patients identified by the feature in the computed AD cohort. For example, in a feature sequentially entailing elements *a* and *b, a*→b, we provided positive predictive values for *a* and *b* individually and then *a*→b. We color-coded sequences in the dashboard to indicate whether a sequential feature is positively or negatively associated with true AD.

### Model evaluations

To evaluate the models, we used metrics of discrimination, namely areas under the receiver operating characteristics curves (ROC) and precision-recall curve receiver (PR) and calibration error, namely the Brier score and root mean square error RMSE. We computed these metrics for each classification task and model on the held-out test testing data curated through chart reviews.

## Results

4,985 patients met our inclusion criteria for the overall cohort, based on having any diagnosis code (AD, other dementia type, or nonspecific dementia) – therefore comprised our Dx AD/Dementia cohort for the first classification task. Nested within this cohort of patients, 1,093 had a specific AD diagnosis record, constituting our Dx AD cohort for the second classification task. Aggregated demographic and clinical characteristics of these patients are provided in Table 1. In the gold-standard data used for testing the algorithms, 33 patients (of the 150 chart-reviewed) were labeled positive for AD. 56 (37.3%) of the chart-reviewed patients had a specific diagnosis code for AD, of whom 32 were labeled positive for AD – therefore, positive predictive value (PPV) for AD diagnosis was 57.1 percent in the Dx AD cohort.

The GTL^16^ algorithm was able to curate labels in the training set that, if tested against the held-out gold standard test data, produced an area under the ROC (AUROC) equal to 0.823. The GTL model used here curated AD phenotype probabilities, in an unlabelled cohort of patients with at least an AD or Dementia diagnosis code, by transferring healthcare utilization patterns learned from 16 diseases: atrial fibrillation, asthma, bipolar disorder, breast cancer, coronary artery disease, crohn’s disease, congestive heart failure, chronic obstructive pulmonary disease, epilepsy, gout, hypertension, rheumatoid arthritis, schizophrenia, stroke, type 1 and 2 diabetes mellitus, and ulcerative colitis. To curate binary labels for the training set, we set the operating point (OP) to 0.1. For consistency, the same OP was used to curate the computed AD cohort from the tSPM features (Table 1).

**Table 1.** Demographic and clinical characteristics of the patient cohorts.

|  | **Dx AD/Dementia cohort** | **Dx AD cohort\*** | **Computed AD cohort\*\*** |
| --- | --- | --- | --- |
| Patients | 4,985 | 1,093 | 663 |
| Mean age | 72.58 | 77.93 | 79.94 |
| Mean Charlson score | 4.57 | 4.75 | 4.59 |
| Mean data depth (years) | 19.49 | 21.58 | 22.23 |
| Mean phenotype record | 3 | 12 | 17 |
| percent inpatient visits | 25 | 13 | 11 |
| percent outpatient visits | 35 | 46 | 49 |
| Unique EHR records | 97,222 | 54,882 | 43,360 |
| percent Female | 47 | 52 | 52 |
| percent African American | 5 | 3 | 3 |
| percent LatinX | 3 | 3 | 2 |
| percent White | 88 | 90 | 90 |

\* Dx AD cohort is a subset of Dx AD/Dementia cohort who has at least a diagnosis record for AD. \*\* Computed AD cohort comprises patients from the Dx AD/Dementia cohort who are likely to have AD based on the tSPM model's predicted probabilities for AD.

### Feature engineering

We mined 162,245,302 transitive sequences from 97,222 unique EHR records from the overall study population of 4,985 patients. 12,501,934 of the sequences passed the sparsity filter in MSMR. The entropy-based feature selections in the MSMR, along with boosting, resulted in a final list of 219 features (Table 3S in the Appendix), which include age and gender as variables.

### Model performances

As demonstrated in Table 2 and Figure 2, the model with tSPM sequences outperformed the model using AD diagnosis codes (Dx AD) in both discrimination and calibration metrics. On the discrimination metrics, the AUROCs and the precision-recall curves were between 2.88 to 9.84 percent improved in the sequential model. The calibration error metrics also showed improvement–between 3.24 to 30 percent in the models with EHR sequences, compared with the model with only AD diagnosis records (Dx AD).

**Table 2.** Model performance metrics

| Classification Task |  | 1: Dx AD/Dementia* |  |  |  | 2: Dx AD** |  |  |  |
| --- | --- | --- | --- | --- | --- | --- | --- | --- | --- |
|  |  | Discrimination |  | Calibration |  | Discrimination |  | Calibration |  |
| model | Metric | ROC | PR | Brier | RMSE | ROC | PR | Brier | RMSE |
| Dx AD*** |  | 0.945 | 0.860 | 0.077 | 0.278 | 0.823 | 0.877 | 0.190 | 0.436 |
| tSPM**** |  | 0.973 | 0.895 | 0.073 | 0.269 | 0.904 | 0.924 | 0.133 | 0.364 |
| improvement |  | 2.88% | 3.91% | 5.19% | 3.24% | 9.84% | 5.36% | 30.0% | 16.51% |

\* classification task 1 classifies AD patients from patients with an AD or Dementia diagnosis code. \*\* classification task 2 classifies AD from patients with at least an AD diagnosis code. \*\*\* GBM model trained with diagnosis codes for AD \*\*\*\* GBM model trained with tSPM sequences

**Figure 2.**
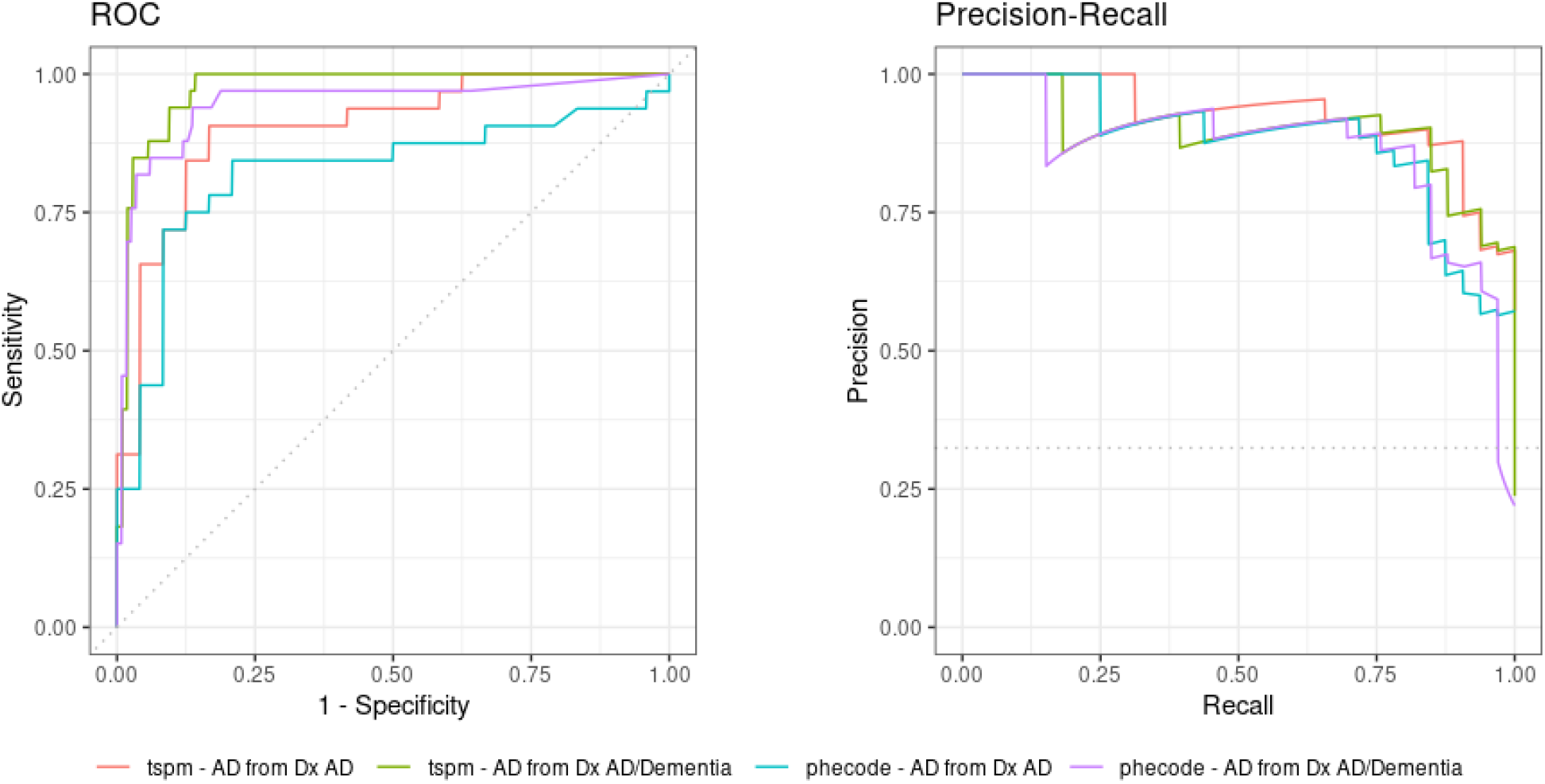
The ROC and PR curves grouped by the task and features. Classification task 1 identified AD patients from patients with an AD or Dementia diagnosis code, whereas classification task 2 identified AD from patients with at least an AD diagnosis code. Phecodes include all diagnosis codes for AD (supplementary table). tSPM models use sequential patterns of medications and diagnoses from the EHR data.

These improvements are, in fact, more substantial than the percent net improvement if considered in the reachability context. For instance, improving AUROC from 0.945 (the Dx AD model in the classification task AD from Dx AD/Dementia) to 0.973 (with the tSPM sequences) represents only 2.88 percent net improvement, whereas the total improvement reachable is about 5.5 percent to reach perfect discrimination (ROC:1). In this context, the tSPM improvement is over 50 percent of the reachable improvement.

Based on our trained tSPM model, 663 patients would be included in the AD cohort. 35 patients in the computed cohort did not have an AD diagnosis code, which suggests that our model is, to some extent, capable of detecting AD undercoding. Compared to the two diagnosis code-based cohorts, patients in this computed cohort were slightly older, on average, had more diagnosis codes for AD and outpatient encounters, and had fewer inpatient encounters (Table 1). Among the demographic variables included in the models, age was the 30th and 10th important feature for classification tasks 1 (classifying true AD patients from patients with AD or Dementia diagnosis code) and 2 (classifying true AD patients from patients with an AD diagnosis code), respectively. Gender was not an important feature for classification in task 1 and marginally important (ranked 63) for task 2.

### Do the identified sequential features make clinical sense?

Among the 219 sequences we found for classifying Alzheimer’s Disease, all diagnosis records that were the first element of the sequence (i.e., *a* in *a*→b) were ICD-9 codes, representing a possibly older record. Except for 2, all diagnosis records that were the later element of the sequence (i.e., *b* in *a*→b) were ICD-10 codes, representing a possibly newer record. We categorized the important sequences under the following groups. Visualizations in this section are provided from an interactive dashboard we developed to visualize sequential pathways and study positive predictive values for sequences and their elements (Figure 1S, Appendix). The positive predictive values are computed based on the true positive patients identified by the feature in the computed AD cohort.

#### 1 A symptom in the past followed by an AD diagnosis or medication

The top 2 important sequences shared between the two classification tasks were sequences of an AD/Dementia symptom followed by an AD diagnosis or medication. An example of such sequences is memory loss (78093 - ICD 9 Diagnosis Code) followed by AD (late-onset G30.1 or unspecified G30.9) ICD-10 code. Memory loss is a common symptom of Alzheimer’s disease. Also, a past record of memory loss ICD-9 code followed by AD medications such as Memantine and donepezil can positively indicate true Alzheimer’s Disease. Our data shows that memory loss alone has a relatively low positive predictive value (∼41%) for truly identifying AD (Figure 3). The PPV for Memantine (5 mg) and donepezil (10 mg) is 56% and 62%, respectively. However, when sequentially paired with memory loss, the respective positive predictive values increase to 71% and 72%.

**Figure 3.**
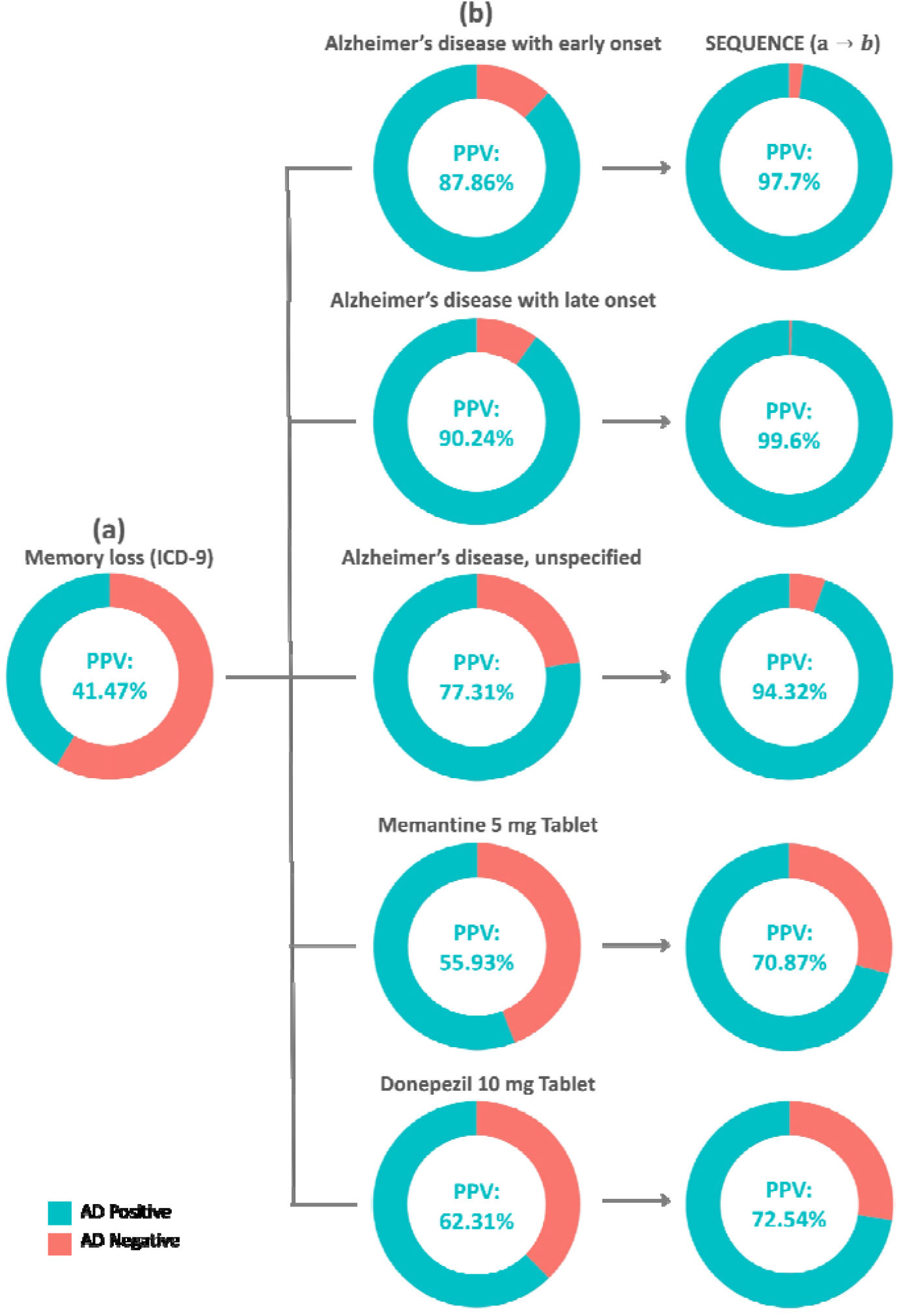
Comparing positive predictive values of the elements of a sequential feature with memory loss for classifying AD.

#### 2 A risk factor in the past followed by an AD diagnosis

The 3rd most important sequential feature in classifying AD encompassed a potential risk factor in the past followed by an AD diagnosis. Notable in this category was unspecified essential hypertension (ICD-9 code 4019), followed by AD ICD-10 diagnosis code for AD or Dementia. Hypertension in midlife is particularly associated with an increased risk of developing dementia and Alzheimer’s disease.^25,26^ Also in this category were past records of unspecified hyperlipidemia (ICD-9 code 2724) or hypercholesterolemia (ICD-9 code 2720), followed by AD diagnosis code (Figure 2S, Appendix).

#### 3 Sequences involving AD medications

Some of the important sequential features we found for classifying AD included sequences of AD medications and diagnosis codes for AD or dementia (Figure 4). Among medications, Donepezil and Memantine were primarily included. For instance, sequences of donepezil with AD or dementia diagnosis records and AD followed by Memantine carry positive signals for classifying AD (Figure 5).

**Figure 4.**
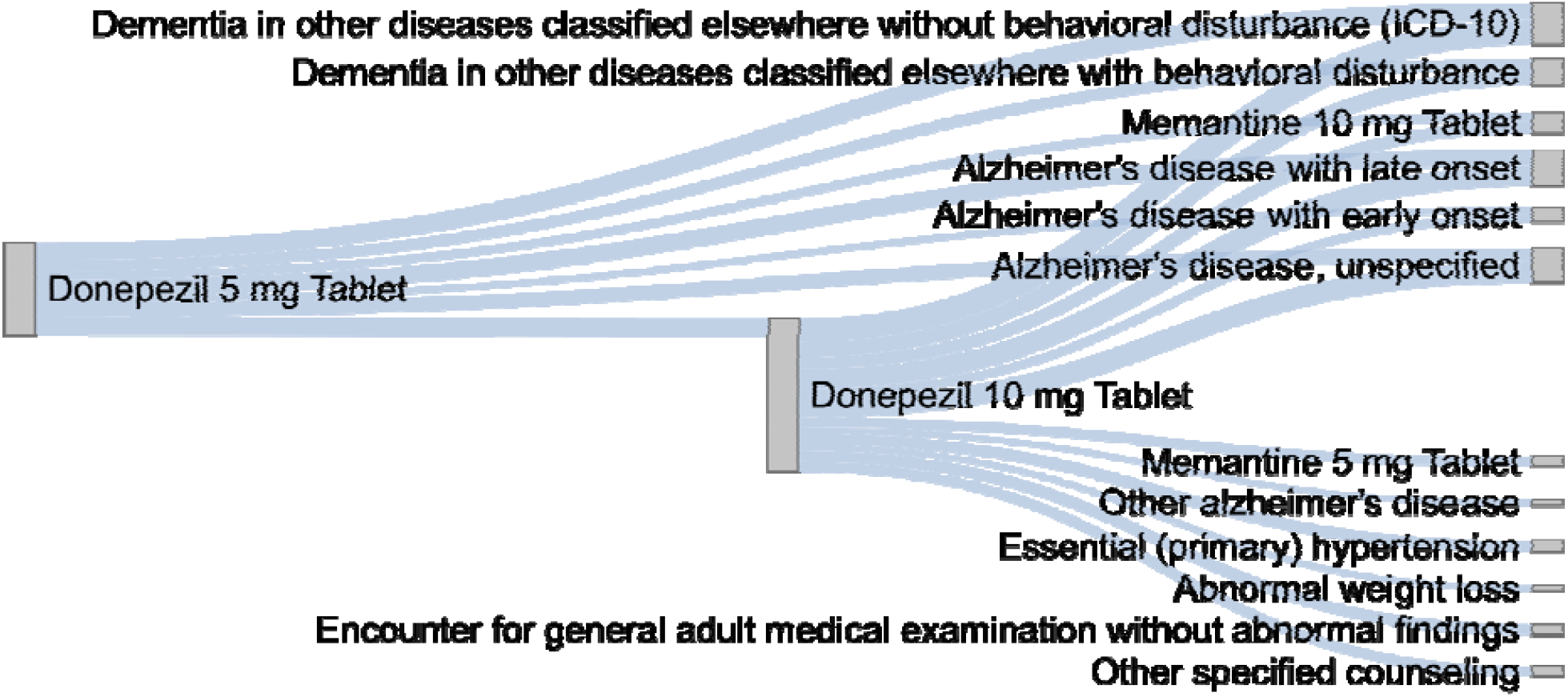
Sequential pathways that indicate true AD compiled by connecting identified sequences of AD medications and diagnoses in the EHR. The visualization dashboard compiles sequential pathways by connecting identified sequences of EHR observations. The pathways presented here involve donepezil and Memantine.

**Figure 5.**
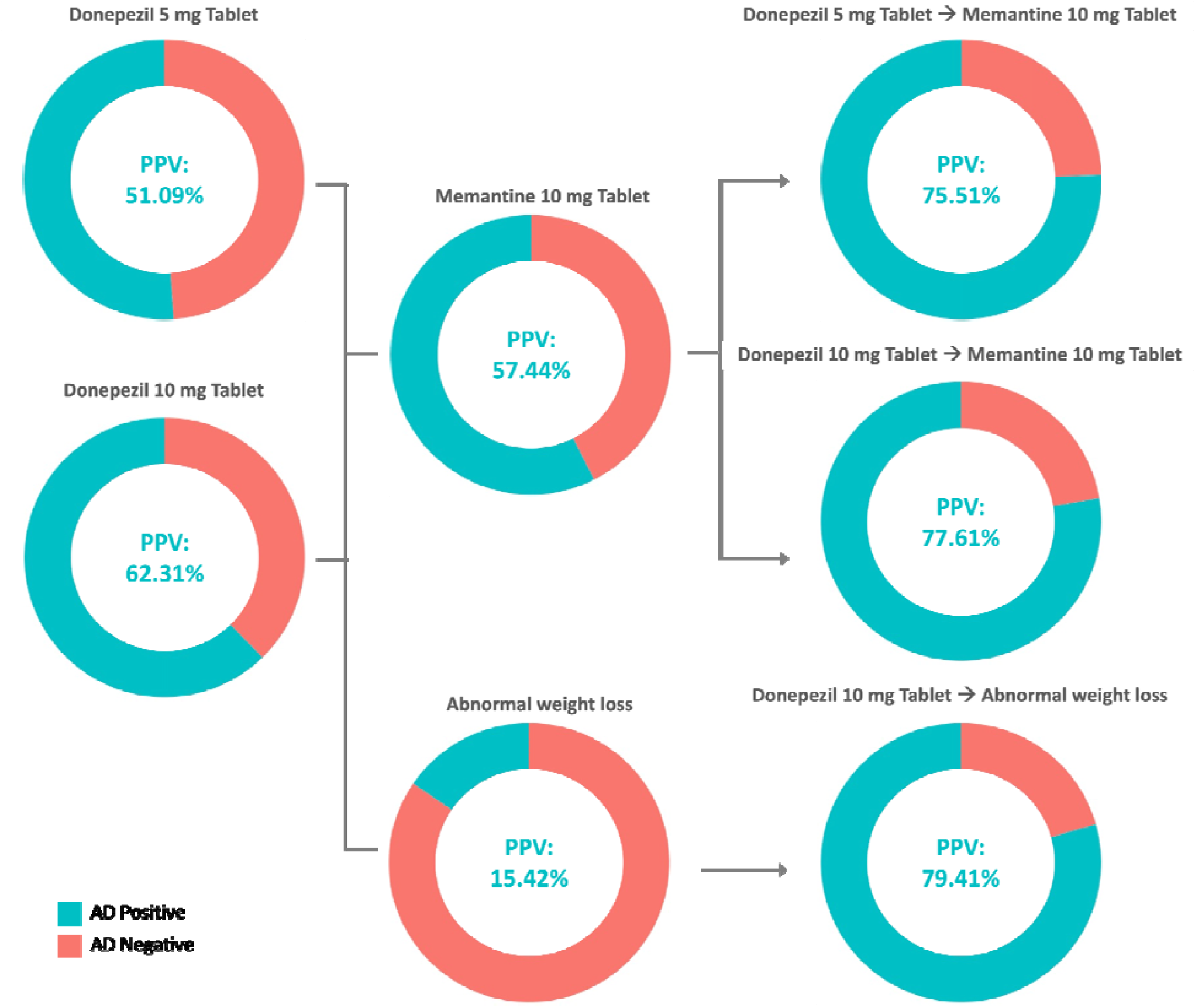
Comparing positive predictive values of the elements of a sequential feature with AD medications that can improve AD classification.

Other important sequences of AD medication reflected changes in medications or in dosage. A highly important sequence, for example, indicated an increase in dosage for donepezil from 5mg to 10 mg. Another sequence possibly reflected a change in the treatment plan from donepezil to Memantine. Donepezil is a medication typically the first medication used to treat Alzheimer’s disease, whereas Memantine is indicated for moderate to severe Alzheimer’s disease, though is sometimes used earlier if the patient cannot tolerate donepezil, or requests it for other reasons.

#### 4 Indirect medication relations to AD diagnosis

We found that the sequence of Sodium Chloride IV followed by Alzheimer’s disease with early onset is an important marker for classifying AD patients. The positive predictive value of the sequence is 96.72 percent, whereas PPV for Alzheimer’s disease with early onset and Sodium chloride IV alone is 87.86 and 9 percent, respectively. Sodium chloride IV solution is commonly used to treat a variety of conditions, including dehydration, electrolyte imbalances, and certain medical emergencies, and is. Several factors can also lead to dehydration in people with AD/dementia, including decreased thirst (or difficulty recognizing thirst), difficulty remembering to drink enough fluids or communicate their thirst, and due to AD or other medications that can lead to dehydration. Some medications used to treat AD, such as cholinesterase inhibitors, can cause side effects such as diarrhea and increased urination, which can contribute to dehydration. Other AD medications, such as memantine, can cause side effects such as constipation, which can also contribute to dehydration if it leads to infrequent bowel movements.

#### 5 Comorbidities with shared roots or as side effects

We found that some sequences of comorbidities with a possible shared cause with AD or AD side effects can potentially indicate true AD. Examples included sequences of different joint pains followed by an AD diagnosis or medication as important digital markers for AD phenotyping (Figure 6 and 3S in the Appendix). Pain in a joint can have a number of causes, including injuries, overuse, or underlying medical conditions, such as Rheumatoid Arthritis (RA), Osteoarthritis (OA), Peripheral arterial disease, and Compression fractures. Both RA and AD diseases are associated with older persons and genetic factors. Besides the inflammation associated with RA, reduced blood flow to vital body organs can increase the risk of developing dementia. Additionally, anti-rheumatic medications used by RA patients may increase the risk of developing dementia.^27^

**Figure 6.**
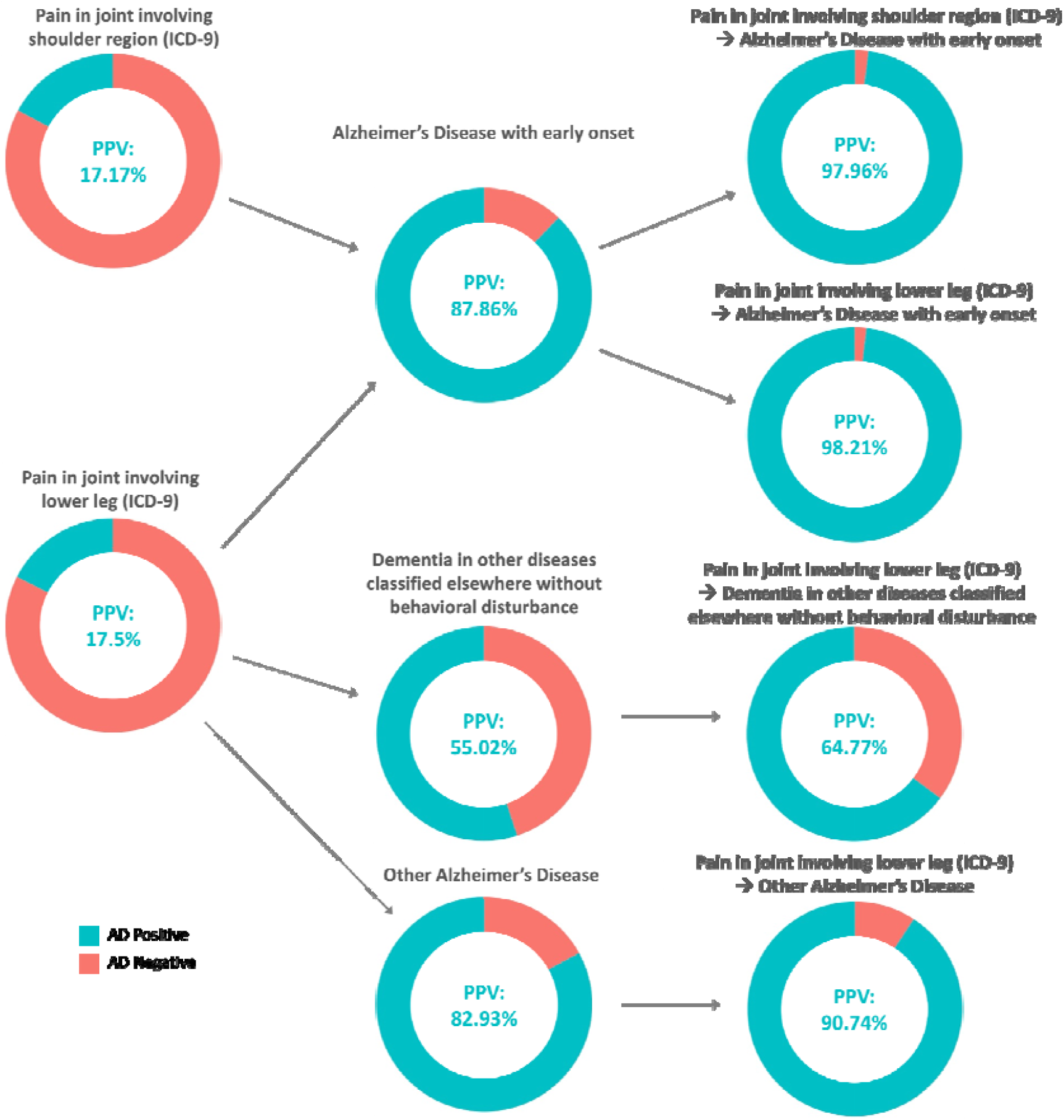
Comparing positive predictive values of the elements of a sequential feature with joint pain for classifying AD.

Another sequence in this category is Anemia unspecified (ICD-9 code), followed by an AD or dementia diagnosis code. Less than 14% of patients in our cohort who have an anemia record are positive for AD. In this setting, an unspecified AD ICD-10 code has a PPV of 77.3 percent. When sequentially paired, Anemia followed by a non-specific AD has a PPV of 81.8 percent. When paired with a late-onset AD diagnosis code, the PPV goes up to 94.3. In both sequences, the positive predictive value for AD increases by about 5 percent. Several studies have found that older adults with anemia may have an increased risk of developing Alzheimer’s disease.^28–30^

#### 6 Repeated AD diagnosis codes

Sequences that embody a temporal repetition of the diagnosis codes are also important in confirming the AD diagnosis. For example, one of the top features we found shows that if a patient has the ICD-9 code 3310 (Alzheimer’s disease) followed by an ICD-10 diagnosis code for AD, chances are higher that the patient truly has AD.

## Discussion

Real-world clinical data stored in the electronic health records systems offer significant opportunities for developing powerful tools to improve epidemiologic evidence on Alzheimer’s Disease at a lower cost than face-to-face clinical studies. However, due to known data reliability issues, study cohorts from EHR data need to be carefully defined in order to minimize the introduction of additional and unnecessary noise in the study. This can be achieved through expert-driven algorithms, which are often costly,^31–33^ or carefully-developed computational models that can scale into digital health tools. Given the projected number of older adults that will develop AD in the future (as they age), scalable computational cohort characterization models can facilitate identifying those who would benefit from effective disease-modifying therapies (DMTs) as they become available.

AI-based digital health tools – such as one built upon the sequential models developed in this study – for improving AD cohort identification with widely available structured clinical data can accelerate precision drug development (e.g., through accelerating clinical trial recruitment) and improve outcomes research (e.g., via reducing the noise in identifying cases and control). However, prior research on the development of prognostic/diagnostic models of AD has had limited incremental predictive value and external validity, or has required cognitive, genetic, and imaging markers that are not routinely collected in clinical care. Imprecise AD cohort identification can impede the recruitment process for clinical trials aimed at evaluating novel therapies undermining the validity of outcomes research.

EHR observations reflect a complex set of processes that thwart their seamless translation into actionable knowledge. Namely, the raw EHR observation data may not be direct indicators of a patient’s “true” health states at different time points, but rather reflect the patients’ interactions with the system, the clinical processes, and the recording processes. EHR observations are also recorded asynchronously across time (i.e., measured at different times and irregularly), which presents foundational challenges for directly applying standard temporal analysis methods. In this paper, we provided a novel way of identifying digital markers, via transitive sequential pattern mining (tSPM), for the identification of AD in EHR data. Our results demonstrated that, given the limited reliability of AD diagnosis codes, sequential pairs of clinical records stored in the EHRs can augment cohort identification of Alzheimer’s Disease cohorts by a magnitude of 3 to 16 percent on a net improvement, over the use of AD diagnosis codes alone.

We categorized the sequences for identifying Alzheimer’s Disease patients into 6 groups. Most important sequences represented (1) a symptom in the past followed by an AD diagnosis or medication (notably involving past records of memory loss and/or mild cognitive impairment), or (2) a risk factor in the past followed by an AD diagnosis, such as unspecified essential hypertension, unspecified hyperlipidemia or hypercholesterolemia. Another group of sequential patterns involving (3) sequences of AD medications, mainly Donepezil and Memantine, also carried positive signals for identifying AD. These sequences likely represented changes in the agent(s) prescribed due to progression of disease or side effectss. We also found sequences as (4) indirect digital markers for AD, such as indirect medication relations to AD diagnosis (e.g., supply of Fluorodeoxyglucose F18, Sodium Chloride IV, Midazolam followed by AD diagnosis). Sodium Chloride IV, PET scan (where Fluorodeoxyglucose F18 is needed), and Midazolam are all typically administered by a healthcare provider in a hospital or clinical setting. A fifth group of sequential features (5) represented comorbidities with possible shared roots or side effects. Sequences of different joint pains followed by an AD diagnosis or medication were important digital markers for AD phenotyping in this group. The final group of sequences was those that (6) represented two different records of AD diagnosis codes.

Similar to all other work based on observational data from electronic health records, this study has limitations. One of those limitations can be attributed to the data quality. We hypothesize that mining temporal representations from raw EHR data may alleviate some of the data quality issues, but we have not rested this hypothesis thoroughly. In addition, data quality issues, such as lack of longitudinal completeness, can be reflected in the sequential patterns mined. In addition, we used an already-enriched population to build our study on, which included all patients with at least a diagnosis record for either Dementia or Alzheimer’s Disease. Therefore, the generalizability of the models developed here to the general population needs further study. Further, although the data used in this study originate from multiple institutions and care settings, the population distribution is geographically limited. Further evaluation is needed to understand possible fluctuations across geographic regions (and thus demographic characteristics) and care settings.

In addition to the superior classification performance and the hypothesis-generating value of the sequential representations as digital markers of disease, they enable temporal story-telling and thus provide tools for explainable Artificial Intelligence (AI). As the clinical utility of complex AI algorithms has been under scrutiny,^34–36^ explainability has become crucial for elevating clinical utility and thereby impact on patient lives. As this method continues to develop, the classification pipeline, with the story-telling capabilities added through interpretive sequences, can be used in data-driven tools that could facilitate diagnostics and cohort characterization across the AD continuum and enhance public health surveillance, targeted screening, and individualize treatment, improve misdiagnosis rate, and enable more precise planning for health care and caregiving resources. Digital health tools built with tSPM representations on real-world clinical data stored in the EHRs offer significant opportunities for high-throughput precision cohort characterization across the AD continuum and at scale and at a lower cost.

## Data Availability

Protected Health Information restrictions apply to the availability of the clinical data here, which were used under IRB approval for use only in the current study. As a result, this dataset is not publicly available. Qualified researchers affiliated with the Mass General Brigham (MGB) may apply for access to this data through the MGB Institutional Review Board.

## Acknowledgement

NA

